# Applying time series analyses on continuous accelerometry data – a clinical example in older adults with and without cognitive impairment

**DOI:** 10.1101/2020.03.24.20042226

**Authors:** Torsten Rackoll, Konrad Neumann, Sven Passmann, Ulrike Grittner, Nadine Külzow, Julia Ladenbauer, Agnes Flöel

## Abstract

**Introduction:** Current analysis approaches of accelerometry data use sum score measures which do not provide insight in activity patterns over 24 hours, and thus do not adequately depict circadian activity patterns. Here, we used a functional approach to analyze accelerometer data that models activity pattern and circadian rhythm. As a test case, we demonstrated its application in patients with mild cognitive impairment (MCI) and age-matched healthy older volunteers (HOV). Moreover, we assessed the impact of chronotype on distribution of activity data.

**Methods:** Data of two studies were pooled for this analysis. Following baseline cognitive assessment participants were provided with accelerometers for seven consecutive days. A function on scalar regression (FoSR) approach was used to analyze 24 hours accelerometer data. In a second step, analyses were controlled for chronotype using the German version of the morningness-eveningness questionnaire (d-MEQ).

**Results:** Information on 47 HOV (mean age 66 SD 6 years) and 13 patients with MCI (mean age 69, SD 8 years) were available for this analysis. MCI patients displayed slightly higher activity in the morning hours as compared to HOV (maximum relative activity at 7:35 am: 75.6%, 95% CI 2.6 to 200.4%, p = 0.031). After controlling for d-MEQ, disturbed activity patterns were found in MCI of intermediate or evening chronotype, compared to HOV, i.e., MCI presented with higher activities in the morning hours (peak at 8:40 am: 357.6%, 95% CI 92.9 to 985.1, *p* < 0.001) and early afternoon hours (peak at 1:40 pm: 401.8%, 95% CI 63.9 to 1436.4, *p* < 0.001).

**Discussion:** Using a novel approach of FoSR, we found timeframes with higher activity levels in MCI patients compared to HOV which were not evident if sum scores of amount of activity were used. In addition, we found that previously described activity patterns as a function of chronotype swere altered in MCI patients, possibly indicating that changes in circadian rhythmicity in neurodegenerative disease are detectable using easy-to-administer accelerometry.

**Clinical Trials:** Effects of Brain Stimulation During Nocturnal Sleep on Memory Consolidation in Patients With Mild Cognitive Impairments, https://clinicaltrials.gov/ct2/show/NCT01782391?term=NCT01782391&rank=1,

ClinicalTrial.gov identifier: NCT01782391

Effects of Brain Stimulation During a Daytime Nap on Memory Consolidation in Patients With Mild Cognitive Impairment,

https://clinicaltrials.gov/ct2/show/NCT01782365?term=NCT01782365&rank=1,

ClinicalTrial.gov identifier: NCT01782365

## Introduction

Activity assessment in natural environment is a widely used approach to determine habitual differences between cohorts (1) and has been used in different disease populations (2,3) or as a predictor for overall mortality (4). Devices to detect rest-activity patterns of body movements include small inertial measurement units (IMUs). These IMUs assess electrical current caused, for example, by acceleration, which is a proxy measure of a body’s movement in space. These so-called accelerometers or actigraphs capture bodily movement with a high precision in time. Still, common accelerometer assessments report aggregated data on number of steps per day or on amount of time spent in high, low or no activity as well as number of bouts without activity (5). The problem of aggregated data is that it reduces acquired information, thereby losing the high dimensionality of continuous 24 hours recordings. Of note, a more holistic view on activity distributions in daily rhythms has been postulated as an emerging topic lately (6). Thus, more detailed assessments of biological rhythms are needed (7).

Measures of circadian organization of activity patterns also have severe drawbacks: They are mostly limited to the amplitude, mesor and acrophase (time of the peak within the fitted 24 h rhythm) of activity which highlights the extremes of daily activity but have a poor resolution in time (8). Changes in circadian activity responses might be subtle so more precise tools are needed to analyze biological rhythms data. Researchers from Johns-Hopkins University (9,10) and Columbia University (11) introduced the concept of functional data analysis (FDA) to accelerometry data. In a first methodological study Goldsmith et al (11) demonstrated that while standard procedures showed inconclusive results, FDA was able to demonstrate differences in distribution of activity pattern over time. This approach thus might be able to detect subtle changes in circadian organization of activity patterns in a variety of disease states, including incipient neurodegenerative disease.

The pathology of Alzheimer’s disease (AD) serves here as a promising clinical example as AD starts years or even decades before first clinical symptoms become apparent (12). Thus, early diagnosis is mandatory for early treatments in order to delay the progression of cognitive decline. Several markers, including disrupted sleep behavior have been described before as a potential screening target (13). Likewise a disruption of circadian rhythm has been proposed (14). Daily rest-activity patterns are hypothesized to be controlled by circadian functions (15,16) which might thus be well-suited to detect early changes in AD pathology. Moreover, such assessments with small IMUs would constitute non-invasive and low-cost approaches that may allow for screening of large cohorts. In addition, such tools could be employed for repeated follow-up in long-term preventive or therapeutic interventions like lifestyle changes or dietary supplementation.

Here, as a clinical example we applied a functional data analysis algorithm on healthy older volunteers (HOV) and patients with mild cognitive impairment (MCI) in a previously acquired data set to describe the circadian rhythm in both groups, and to test if their activity patterns differed throughout a 24 hours cycle. We hypothesized that the distribution of activity would differ with regard to timing and magnitude between HOV and MCI. In a second step, based on previous reports showing that circadian activity is highly influenced by chronotypes (17), we asked if and how chronotypes (morningness vs. eveningness-types) would modify activity patterns between HOV and MCI.

## Methods

Data reported here were taken from baseline measurements of two intervention studies that assessed the effects of oscillatory tDCS during sleep (daytime nap vs night sleep) on cognitive performance (Effects of Brain Stimulation During a Daytime Nap on Memory Consolidation (study 1) & Effects of Brain Stimulation During Nocturnal Sleep on Memory Consolidation (study 2) in young and older healthy subjects and subjects with mild cognitive impairment (MCI)). These studies are described in detail elsewhere (18,19) and included comprehensive assessment of subjective (using sleep diary and sleep questionnaires) and objective sleep-wake behavior among others, as well as neuropsychological testing and structural imaging of the brain using magnetic resonance imaging (MRI). Objective sleep assessment included seven days of accelerometry. In the current analysis we used all available baseline data of patients with MCI and age-matched HOV.

### Study approval

The studies were approved by the institutional review board of the Charité Universitätsmedizin Berlin, Germany (EA1_295_12 & EA1_028_12), and were conducted in accordance with the declaration of Helsinki (Version 2008). All participants gave written informed consent prior to participation, and received a small reimbursement for their time.

### Participants

HOV (50 – 90 years) were recruited via advertisements in the local database of the Charité Universitätsmedizin Berlin, Germany. MCI patients (50-90 years) were referred to the study from the memory clinic of the Charité University Hospital. The original studies included also young healthy participants; for the current analyses we focused on HOV and MCI, given our aim to compare circadian organization of activity in older adults. Participants underwent a structured telephone interview to exclude the presence of manifest sleep disturbances, contraindications for MRI, and non-native German speakers. Out of 242 potential participants that had entered the study, 66 HOV (mean age 66, SD 6) and 31 MCI (mean age 70, SD 8) were then invited to the laboratory to determine study eligibility which included a clinical interview, neurological examination, structural MRI, and standardized cognitive testing. Inclusion and exclusion criteria for both groups are detailed in the supplementary. In short HOV participants had to show no signs of dementia, and no present episode of depression (monitored with the Beck’s Depression Inventory (BDI (20))) while MCI patients had to fulfill core clinical criteria for the diagnosis of MCI outlined by Petersen and others (21). Further, clinical assessment and structural MRI revealed no systemic or brain diseases accounting for declined cognition. Patients diagnosed with amnestic or amnestic plus MCI were included.

### Neuropsychological Assessment

Both groups were assessed using a neuropsychological test battery addressing various cognitive functions to ensure that HOV performed within age and gender matched normal range. A detailed description of all domains and tests can be found in the supplementary material (Table S2).

In addition we determined the affective state using the Positive and Negative Affect Schedule (PANAS (22)), anxiety state using the State Trait Anxiety Inventory (STAI-G X1 (23)), self-reported well-being using the World Health Organization Quality of Life-questionnaire (WHOQoL (24)), and the ability to cope with stressful events with a 120-items questionnaire (Stressverarbeitungsfragebogen, SVF (25)) which measured the individual tendency of using specific strategies of stress coping in general (trait). The SVF differentiates 10 strategies aiming on reduction of stress summarized to the following three positive strategies: Positive 1 (Pos 1; including cognitive strategies like reevaluation), Positive 2 (Pos 2, involving distraction-like strategies) and Positive 3 (Pos 3, focusing on control of stress and stress reactions). Moreover, the SVF cover seven strategies which were merged to “negative strategies” (Neg) and are assumed to cause an increase in stress (e.g., rumination, resignation). Lastly, questionnaires were administered to assess subjective sleep habits with the German version of the Pittsburgh Sleep Quality Index (PSQI (26)), daytime sleepiness with the German version of the Epworth Sleepiness Scale (ESS (27)), and individual chronotypes with the German version of Morningness–Eveningness-Questionnaire (d-MEQ (28)).

### Accelerometry

The ActiGraph GT3X+ (ActiGraph, Pensacola, FL, USA) was used in this study. It is able to assess acceleration in the vertical, antero-posterior and medio-lateral axes. It has shown high inter-instrument reliability (Intraclass correlation 0.97 (29)) and intra-instrument reliability within frequencies that are common in human activities, and is described as a reasonable tool for longitudinally measuring sleep (30). During the week following baseline cognitive assessment each participant wore a GT3X+ on the hip (figure S2) for continuous seven-day recordings to fully capture daily physical activity. Subjects were asked to wear the device at all times and just to remove it for any activity involving water (showering, swimming, etc). The devices were pre-programmed with default settings (30 Hz in three axis, with a fixed start and stop time). Data were downloaded in 60 sec epochs. Accelerometry data download and sum score descriptives were performed using ActiLife Software 6.8.2 (ActiGraph, Pensacola, FL, USA).

After download, data were first checked for its actual wear time. The default algorithm according to Choi (31) was used to identify non-wear time. Days of wear time of < 60 % were excluded from further analysis. Raw values of accelerometry assessment are changes in current which are conversed to activity counts (32). Activity counts are an abstract value, usually used in literature. Even though they constitute an abstract value derived from company owned algorithms, higher values represent more intense activity compared to lower values. Different cut-offs to classify activity are available. We picked 0 – 99 activity counts per minute as sedentary activity, 100 – 2019 as light activity, 2020 – 5998 as moderate and everything above 5999 as vigorous activity (33) as these cut-points derived from a large NHANES cohort of 6329 participants and thus were assumed to correctly reflect levels of activity.

### Statistical Analyses

We applied a Function-on-Scalar-Regression (FoSR) approach that is in line with recent publications by Goldsmith et al (11) and the group of Biostatistics from Johns Hopkins University (9,10,34). Function on scalar in this study means that activity counts as main outcome are functions of time (response), while covariates, here MCI patients/HOV etc. are scalars. However, for our analysis we made two important modifications:

– The set of basis functions used in the regression model are discrete wavelet functions instead of cubic splines (7).
– In addition to the fixed effects, the model equation contains a random intercept to account for clustering of data within individuals.

Pre-processing steps for FoSR as well as all statistical analyses were performed using statistical software R (Version 3.2.1) and the R package ‘wavelets’ (version 0.3-0) (35).

For FDA activity count data needed to be aggregated to at least 5 min epochs according to Xiao et al (10) resulting in 288 count data sets per participant per day (24h=1440 minutes=288*5 minutes). Furthermore, we log-transformed activity counts as distribution of count data were skewed. For high resolution accelerometer data robustness of data is a problem as changes between zero activity and high activity peaks might be present within small time periods. Furthermore, rapid changes in activity might not reflect circadian rhythm activity patterns. Therefore, a smoothing algorithm is needed to distinguish between signal (circadian activity pattern) and noise (changes between zero activity and high activity peaks within small time periods). Cubic splines or wavelet transformation are both suggested for smoothing of this type of data. Discrete wavelet functions and cubic splines have both a localized support, meaning that each wavelet function / each piece of the cubic spline model is able to fit the activity data well in a particular time range and reduces noise in this time range. In contrast to cubic splines, wavelets are periodic basis functions (with a period of 24 hours) and are thus better suited for modelling circadian patterns. Here discrete wavelet transformation was used for smoothing activity counts.

### Discrete Wavelet Analysis

The discrete wavelet transformation is a well-established method in the analysis of time series (c.f. (36) and (37) pp. 174-179). Different series of wavelets are in use for the analysis of time-series (Coiflets, Least Asymetric, Best Localized, Daubechies wavelets, all implemented in the R package ‘wavelets’). The full discrete wavelet transformation is a regular linear (matrix) transformation from a finite dimensional real vector space into a space of the same dimension. Here, we used incomplete wavelet transformation with a non-trivial kernel that maps the time series from a high dimensional space (dim=288) into a real space with far less dimensions (dim=18, 18 basis functions). The purpose of this approach is twofold:

– Smoothing the activity data (removing noise).
– Defining a set of basis functions which can be used in the function-on-scalar regression analysis for testing group/subgroup specific activity patterns.

The incomplete discrete wavelet transformation behaves like a low-pass filter, meaning that rapid changes between non-activity and high activity will be smoothed out. For our analysis, we chose Daubechies wavelets of length 10 (d10, which corresponds to a time range of 50 minutes for the first level wavelet functions), meaning that an activity pattern of 50 minutes is modelled by one function.

The degree of smoothing depends on the number of basis functions used, which results from the length of the functions. Shorter wavelet filters tend to give poor results in smoothing the data (not enough smoothing, too much noise) whereas wavelets with long filter length are not well localized (too much smoothing, no sufficient model fit with regard to pattern relevant changes in activity). The use of 18 basis functions seemed to be a good compromise between good smoothing results and preserving a sufficiently high resolution for detecting activity patterns within circadian rhythm. The incomplete wavelet transform in concise matrix notation is

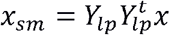

*x* denotes the vector containing the raw data whereas *x*_*sm*_ is the vector of smoothed data (the subscript *sm* stands for “smoothed”). The columns of the matrix *Y*_*lp*_ are the 18 basis function of the incomplete wavelet transform. Since each basis function is a vector of length 288, the matrix *Y*_*lp*_ has 288 rows and 18 columns. Finally, the subscript *lp* refers to the low pass property of the incomplete wavelet transform.

### Function-on-Scalar-Regression model

The presentation of the technical details of Function-on-Scalar-Regression closely follows the presentation in the appendix of (36). The model equation of FoSR is largely analogous to the model equation of a linear mixed effects model. We included a random intercept in the model equation since some participants contributed several correlated 24 hours activity records to the study.

Let *x*_*i,j*_ be the vector of a log(1+count) transformed 24 hour activity record for day *j* = 1,…, *j*_*i*_ of participant *i* =1,…,N aggregated in 5 minutes time blocks. Since each volunteer wore the actigraph device between one and eight days, *j*_*i*_ may range from 1 to 8. Hence, the model equation is

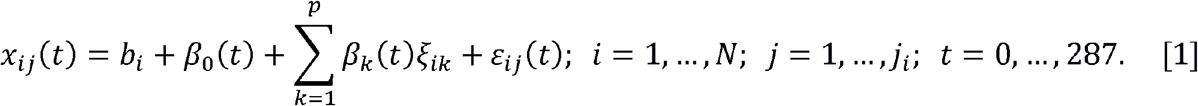

In equation [1] ξ _*ik*_ enotes the value of the *k*^th^ covariate of the *i*^th^ participant and *ε*_*i,j*_ (*t*) are the normally distributed residual terms with expectation zero and common variance *σ*^2^. The random intercept terms *b*_*i*_ for the participants are normally distributed with expectation zero and variance *τ*^2^. We expanded each (row) coefficient vectors *β*_k_(*t*) using the wavelet basis functions that are the columns of *Y*_*lp*_

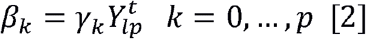

Omitting the index *t*=0,…, 287, the vector *β* _*k*_ has length 288 and γ_*k*_ is a row vector of length 18 (number of wavelet basis functions). Details of the estimation of model parameters are presented in the appendix.

The FoSR model provides for each covariate (e.g. a group indicator variable) absolute and relative differences of activity counts with 95% confidence intervals and p-value for each five minutes époque. We report periods where the covariate (e.g. an indicator variable defining group differences) is significant in all five minutes époques and report only the largest relative difference in percent with two-sided 95% CI and p-value. All analyses were controlled only for age in order not to overfit the model.

### Addressing the problem of multiple testing

All p-values were Bonferroni adjusted by a factor of 18. The Bonferroni factor was chosen equal to the number of basis functions used in the FoSR model. Accordingly, the confidence level of the confidence bands was risen from 1-α to 1-α /18.

### Secondary analysis

For our secondary analysis we focused on the impact of chronotypes on the circadian organization of activity within both groups. We therefore stratified the analysis by chronotype groups: morning (moderate and definite morning types) and intermediate/evening chronotypes (intermediate, moderate, definite evening types) according to d-MEQ. We compared activity patterns of MCI and HOV within each strata.

## Results

97 HOV and MCI patients were available for inclusion (patient flow is displayed in Figure S1 of the appendix). Of these 97 participants, 37 had to be excluded either due to missing actigraph data, abnormal MRI findings that suggested a differential diagnosis (e. g., stroke), or due to insufficient wear time of the actigraph (19 HOV, 18 MCI patients). An additional four MCI patients had to be excluded due to incoherent zero values distributed over a 24 hours cycle. Thus 47 HOV (mean age 66, SD 6) and 13 MCIs (mean age 69, SD 8) remained for final analysis with 228 valid accelerometer days in HOV and 60 days in MCI respectively. We compared basic characteristics of included and excluded subjects but could not identify major differences.

### Baseline characteristics

Information on baseline characteristics including sociodemographic characteristics and cognition can be found in Table 1. Overall, MCI patients differed from HOV with regard to age, (mean 69 vs. 65 years, SMD = 0.51), depressive symptoms (median 7 vs. 3 in BDI, SMD=0.7), cognitive functioning (mean MMSE 28 vs. 29, SMD=1.07) and in positive and negative stress coping strategies (lower values in MCI with SMD from 0.99 to 1.39). The d-MEQ displayed higher rates of morning types in MCI compared to OLD (moderate morning type: n=17 (37.0%) vs. n=6 (46.2%); definite morning type: n=6 (13.0%) vs. n=3 (23.1), SMD=0.53) while subjective sleep ratings did not show substantial differences (median 4 vs. 3 in PSQI, SMD=0.24).

**Table 1:** Baseline characteristics and aggregated descriptive accelerometer data for HOV and MCI.

|  | HOV<br>(n = 47) | MCIs<br>(n = 13) | SMD<br>standardized<br>mean difference |
| --- | --- | --- | --- |
| Female sex, no (%) | 25 (53.2) | 8 (61.5) | 0.17 |
| Age in years, mean (SD) | 65 (6) | 69 (8) | 0.51 |
| Education in years, mean (SD) | 16 (3) | 15 (3) | 0.30 |
| MMSE, mean (SD) § | 29 (1) | 28 (2)* | 1.07 |
| Becks Depression Inventory, median (IQR) \| | 3 (1 – 4) | 7 (4 - 11)* | 0.70 |
| STAIG-X1, mean (SD) \| | 33 (10) | 37 (9) | 0.40 |
| WHOQoL overall score, mean (SD) | 100 (14) | 93 (19) | 0.47 |
| PANAS positive trait, median (IQR) | 3 (3 – 4) | 3 (3 – 4) | 0.50 |
| PANAS negative trait, median (IQR) | 1 (1 – 1) | 1 (1 – 1) | 0.42 |
| SVF positive strategy 1, mean (SD) # | 12 (3) | 8 (6)* | 0.99 |
| SVF positive strategy 2, mean (SD) # | 12 (3) | 7 (5)* | 1.23 |
| SVF positive strategy 3, mean (SD) # | 17 (4) | 10 (7)* | 1.18 |
| SVF negative strategy, mean (SD) # | 9 (3) | 4 (4)* | 1.39 |
| PSQI sum score, median (IQR) ¶ | 4 (3 – 5) | 3 (3 – 5) | 0.24 |
| ESS, mean (SD) † | 7 (4) | 6 (3) | 0.16 |
| d-MEQ, no (%) |  |  | 0.53 |
| definite evening type | 1 (2.2) | 0 (0.0) |  |
| moderate evening type | 2 (4.3) | 1 (7.7) |  |
| intermediate type | 20 (43.5) | 3 (23.1) |  |
| moderate morning type | 17 (37.0) | 6 (46.2) |  |
| definite morning type | 6 (13.0) | 3 (23.1) |  |
| **Sleep scoring** | **HOV** | **MCI** | **SMD** |
|  | (n = 47) | (n = 13) | **standardized**<br>**mean difference** |
| Time in bed, mean (SD) | 00:19 (58) | 00:00 (49) | 0.00 |
| Time out of bed, mean (SD) | 08:40 (69) | 08:20 (25) | 0.20 |
| Total time in bed (min), mean (SD) | 496 (62) | 495 (84) | 0.01 |
| Total sleep time (min), mean (SD) | 483 (63) | 479 (79) | 0.06 |
| Wake After Sleep Onset (%), mean (SD) | 2.4 (1.5) | 2.9 (1.6) | 0.42 |
| Arousals (no.), mean (SD) | 4.1 (2.1) | 4.4 (2.2) | 0.12 |
| Average Awakening (min), mean (SD) | 3.1 (0.9) | 3.5 (1.0) | 0.40 |
| Sleep latency (min), mean (SD) | 1.7 (1.0) | 1.6 (1.0) | 0.07 |
| **Physical activity** | **HOV** | **MCI** | **SMD** |
|  | (n = 47) | (n = 13) | **standardized**<br>**mean difference** |
| Energy Expenditure (kcal/day), mean (SD) | 417 (175) | 368 (303) | 0.20 |
| Time in sedentary activity (%), mean (SD) | 74.1 (7.6) | 72.8 (8.0) | 0.17 |
| Time in light activity (%), mean (SD) | 22.8 (6.4) | 24.5 (5.5) | 0.30 |
| Time in moderate activity (%), mean (SD) | 2.5 (1.3) | 2.7 (3.5) | 0.09 |
| Time in vigorous activity (%), mean (SD) | 0.1 (0.3) | 0.0 (0.0) | 0.49 |
| Moderate-to-vigorous physical activity (%), mean (SD) | 2.6 (1.6) | 2.6 (3.5) | 0.04 |

§ MMSE denotes mini mental state examination and is a score ranging from 0 to 30 screening for overall cognitive functioning in which higher values describe better cognitive functioning.
| 4 missing at random (3 in HOV, 1 in MCI)

Participants spent most of their day in a sedentary lifestyle or in light activity with only small amounts of time in higher activity levels (Table 1), lacking substantial differences between groups (SMD for all measures <0.5). Aggregated analyses of sleep assessed via accelerometry exhibited comparable characteristics between the two groups (most SMD 0.2 or lower) only for wake after sleep onset and average awakening MCIs had somewhat higher values (SMD=0.42/0.40).

### Function-on-Scalar-Regression

We compared the distribution of activity in MCI patients with HOV over the averaged 24 hours day-night cycle using FoSR. Variability of activity levels was higher in MCI patients compared to HOV. Fig 1a displays the smoothed average absolute activity over the course of a 24 hours cycle. MCI showed higher activity levels throughout the waking hours with a slight decrease below levels of HOV in the evening hours. For direct comparison between groups we analyzed relative activity between MCI and HOV adjusted for age (Fig 1b). We observed higher relative activity in MCI patients compared to HOV in the morning hours between 7:25 and 8:00 am peaking at 7:35 am (75.6%, 95% CI 2.6 to 200.4%, p = 0.031). In addition, during the day between 2:35 and 5:05 pm slightly higher level of activity with a peak at 4:10 pm (48.5%, 95% CI −11.7 to 149.6%, *p* = 0.412) was detected in MCI patients compared to HOV. In the evening hours between 8:35 and 10:05 pm MCI patients showed a slightly relative lower activity with its lowest value at 9:30 pm compared to HOV (−34.1%, 95% CI −60.8 to 10.7, *p* = 0.294). Over the course of the sleeping hours (mean time to bed and time out of bed hours in each group are displayed in Table 1) activity patterns of MCI patients and HOV were comparable. In sum MCI patients were more active in the morning hours and activity in MCI had higher variance compared to HOV.

**Figure 1:**
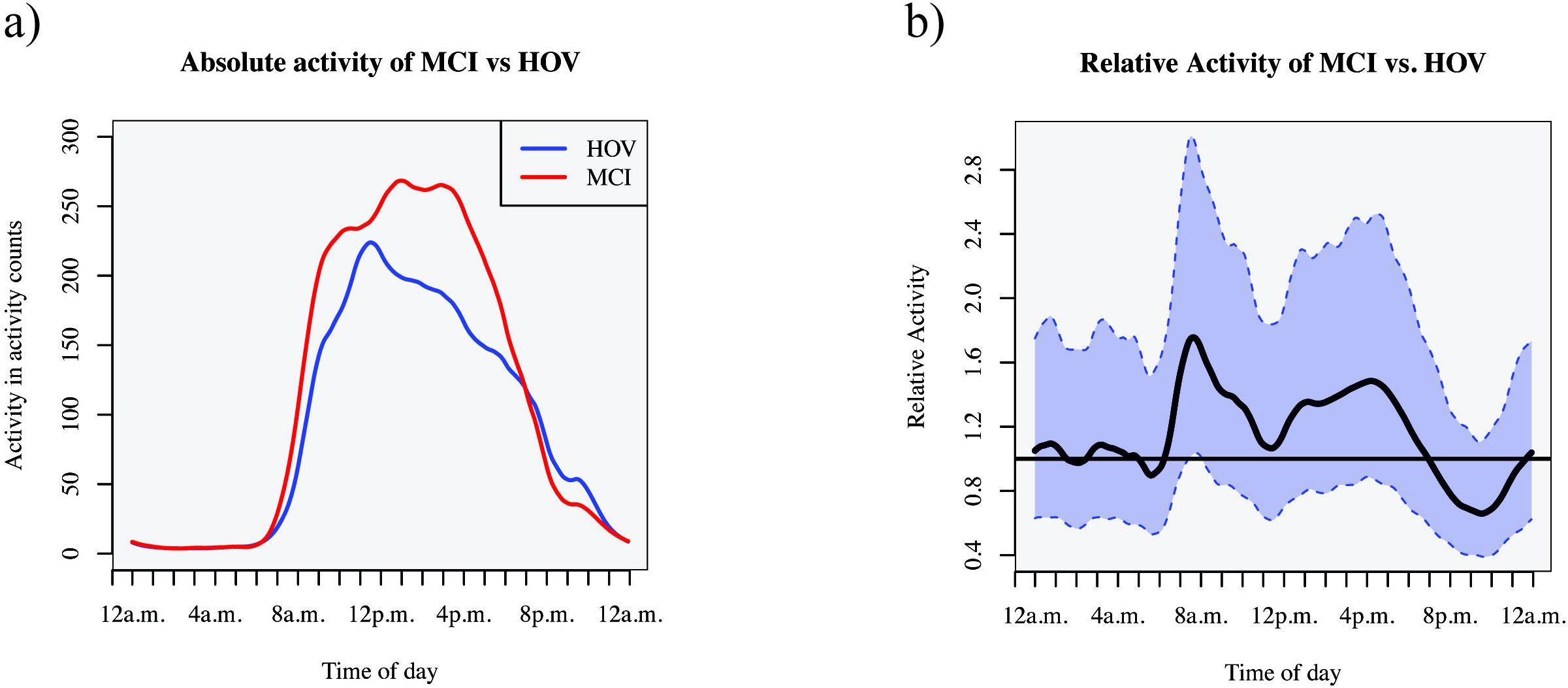
Comparison of absolute and relative activity distribution between MCI and HOV. Distribution of a) mean absolute activity between MCI and HOV over time and b) mean relative activity between MCI and HOV over time with 95% Confidence Interval (CI) adjusted for age.

### Chronotypes

According to the d-MEQ, the majority of participants in the HOV group were intermediate types (44%) while in MCI patients the majority was of moderate morning type (46%) (Table 1).

The HOV group showed the expected pattern of activity with regard to questionnaire-based chronotypes. Compared to moderate and definite morning types combined, the group of intermediate, moderate evening and definite evening types combined displayed higher activity levels in the evening hours between 8:25 pm and 0:20 am with a peak at 11:20 pm (144.4%, 95% CI 48.2 to 303.0, *p* < 0.001) and lower activity in the morning hours between 6:20 and 9:30 am with the lowest activity at 7:50 am (−60.1%, 95% CI −75.2 to −35.8, *p* < 0.001) (Fig 2a).

**Figure 2:**
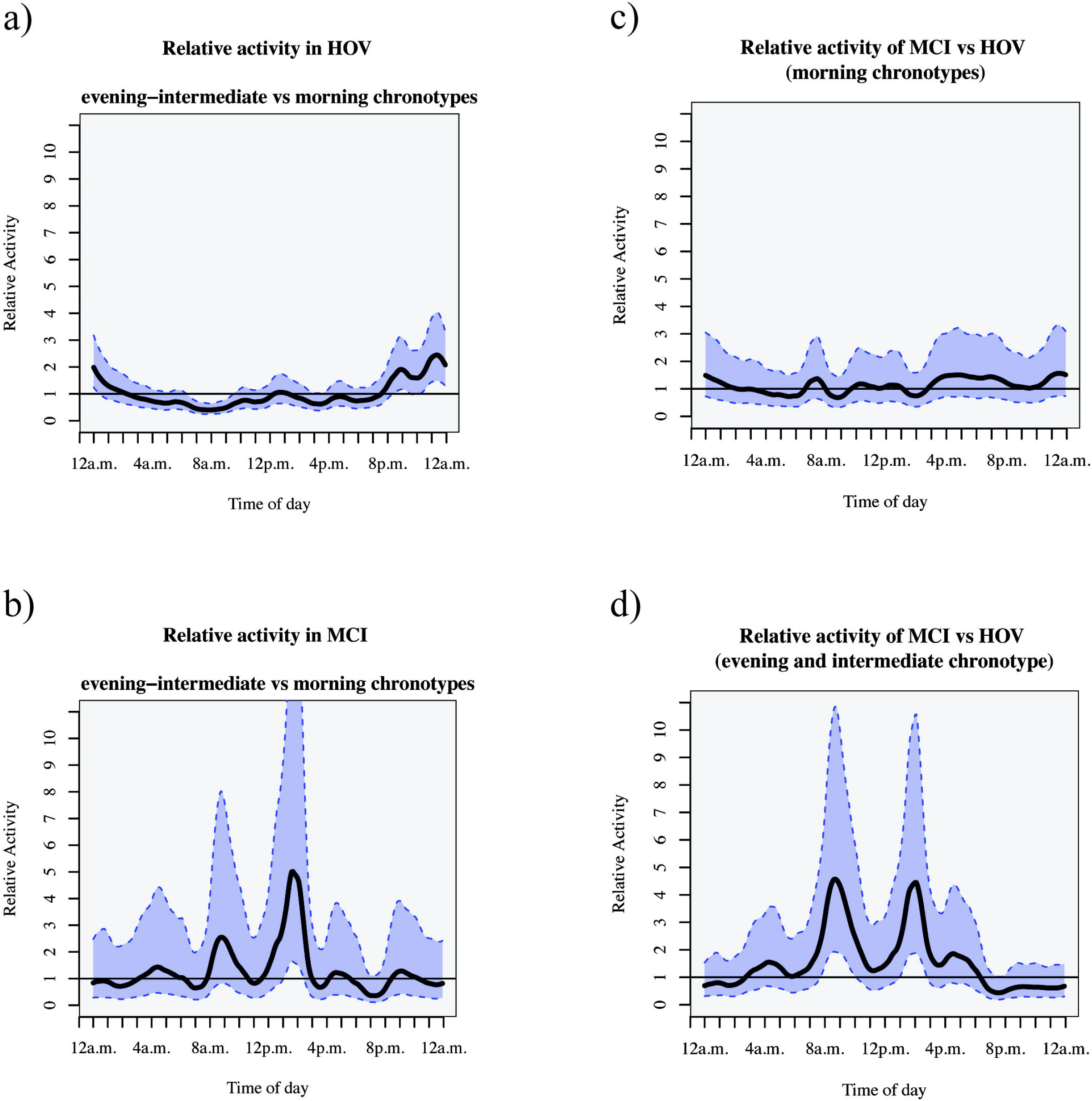
Comparison of activity distribution for different chronotypes. Distribution of mean relative activity over time clustered for the different chronotypes (assessed with the D-MEQ) a) definite evening, moderate evening, and intermediate type vs moderate morning, and definite morning types in MCI, b) definite evening, moderate evening, and intermediate type vs moderate morning, and definite morning types in HOV, c) mean relative activity over time between MCI and HOV for clustered intermediate and evening types, and d) mean relative activity over time between MCI and HOV for morning types. Data are shown with 95% confidence intervals. All analyses are adjusted for age.

In the MCI group definite evening chronotypes were not present. We opposed moderate and definite morning types to clustered intermediate, and moderate evening types as done before (38). Over the course of the day only early afternoon hours between 1:05 and 2:25 pm displayed differences with higher levels of activity in evening and intermediate types peaking at 1:40 pm (401.8%, 95% CI 63.9 to 1436.4, *p* < 0.001) compared to morning types (Fig 2b).

Under the assumption that activity patterns are linked to chronotypes, we would expect no intergroup differences (between MCI and HOV) within specific chronotypes. However, since circadian rhythm might be disrupted in MCI participants, we hypothesized to find group differences within chronotype strata. Subsequently we evaluated intergroup comparison between MCI and HOV for morning chronotypes as well as for evening and intermediate chronotypes separately. While activity patterns in morning types were comparable between groups (Fig 2c) combined groups of intermediate and evening types showed disrupted activity patterns in the morning hours between 7:45 and 10:00 am with a higher peak at 8:40 am (357.6%, 95% CI 92.9 to 985.1, *p* < 0.001) in MCI vs. HOV and early afternoon hours between 1:00 and 2:25 pm with a peak at 2 pm (345.2%, 95% CI 87.7 to 955.8, *p* < 0.001) with higher activity levels in the MCI group (Fig 2d).

To conclude we identified disrupted activity patterns in MCI compared to HOV in the combined group of intermediate and evening chronotypes that were not discernable before this stratified analysis by chronotype.

## Discussion

In this study our goal was to apply the previously published Function-on-Scalar-Regression (FoSR) algorithm for the analysis of accelerometry data to a clinical population to provide an example for the algorithm’s potential in observational or intervention studies. We analyzed data from two pooled, previously published trials with the commonly applied approach in accelerometry, i.e., accelerometer derived sum scores (5,39). Here, we did not find substantial differences in proportions of time spent in specific activity levels or in total volume of activity. Next, we administered FoSR to investigate the circadian organization of activity patterns in patients with MCI and HOV using time series analysis and found higher relative activity in the morning and early afternoon hours and lower activity levels in the evening hours in MCI patients compared to HOV. While the net difference in amount of activity is likely to be negligible, our approach allowed us to demonstrate different time-based distributions of activity between the two populations.

Addressing circadian organization of activity, several authors have used five or ten hours spent in lowest or highest activity to assess phase shifts in activity in patients with MCI or dementia (16,40,41). Overall, findings from these studies have not been unequivocal, which indicates that especially in the early stages of cognitive impairment changes are subtle. Musiek et al. (16) described in their study differential activity patterns distributed throughout the day with lower amplitude between lowest and highest amount of activity and an increased variability over the course of 24 hours in preclinical AD patients. The authors compared different methods to analyze accelerometer data but did not apply time sensitive functional analysis. In contrast, time sensitive functional analysis was central in our study to identify time zones during which MCI patients were more or less active compared to HOV.

In a second step we controlled our regression model for circadian predisposition. We found that MCI patients with either intermediate or evening chronotype showed higher amounts of activity in the early afternoon hours compared to morning types. Moreover, we demonstrated that distribution of activity during a 24-hours cycle was different in intermediate and evening types from activity patterns found in HOV in the present study, or previous reports (38). To the best of our knowledge altered activity patterns in patients with MCI mediated by chronotype have not been described before. Individuals with the eveningness type have been reported to show higher risk of depressive symptoms (42) or insomnia (43), both independent risk factors for cognitive decline. Moreover, a disruption of circadian activity synchrony has been demonstrated in mice models of AD (44,45) and been described in human AD cohorts (46,47). Still contribution of diurnal predisposition on health outcomes is a relatively understudied field especially in AD. Even though our study cannot determine if the differences found between HOV and patients with MCI would lead to earlier detection of MCI in screening of individuals in the community, but this issue can now be addressed in future large-scale studies.

## Limitations

Several limitations should be considered when interpreting our findings. First, groups of HOV and MCI patients differed slightly with regard to age, which might introduce a bias due to altered sleep behavior in the process of aging. However, we believe that these differences did not substantially impact our main results, given that the analysis was controlled for age. Second, we did not adjust for sex in our regression model to avoid overfitting in our small sample. Future studies with larger cohorts should adjust for sex differences to address the issue of potential sex differences. Third, the sample of our MCI group was small. However, we used our data as a test case to demonstrate an analysis approach with potential in a wide range of future applications. The validity of this approach has to be confirmed or refuted in future larger trials. Fourth, we cannot determine from the present data if disturbed activity patterns in MCI patients with the eveningness chronotypes are causally related to the cognitive impairment. Also, morningness types did not show altered activity patterns. However, our results may now be used as a starting point to generate new hypotheses, giving the robustness of the results.

## Future research

First, to better differentiate changes in activity patterns due to chronotypes replication in large longitudinal studies are needed to corroborate our results and confirm the FoSR algorithm as a clinical measure. Also assessment in different pathological and healthy conditions are warranted to extract specific alterations in activity patterns for a certain pathology like AD. Second, results derived from accelerometers with FoSR approach should be compared to wrist-worn accelerometers, to facilitate studies of larger cohorts with commercially available activity monitors. Third, to inform inferences on the underlying mechanisms, biological markers such as melatonin and cortisol secretion or body temperature should be determined to more accurately define circadian rhythms. Finally, it should be assessed if parameters described here are sensitive to changes over time in response to therapies.

## Conclusion

Using a novel approach of FoSR, we were able to demonstrate improved precision in analysis of time based high dimensional activity data. We found that MCI patients exhibited increased relative activity in the morning hours compared to HOV. In addition, we found that previously described activity patterns as a function of chronotype were altered in MCI patients of the eveningness chronotype. The approach of FoSR to analyze activity patterns may constitute an important screening tool for incipient neurodegeneration in the population at large, particularly if accelerometers of commercial smart watches are found to provide with valid measures for FoSR.

## Data Availability

Data and analysis scripts are going to be available with publication at https://doi.org/10.5281/zenodo.3718578

## Acknowledgements

This work was supported by grants from the Deutsche Forschungsgemeinschaft (Fl 379-10/1; Fl 379-11/1, and DFG-Exc 257). We thank Lena Reich for their help with data acquisition. Special thanks to Patrizia Müller and Madlee Einsiedler for data preparation and actigraphy sleep ratings. Additionally, we want to thank Luo Xiao, Jeff Goldsmith and Ciprian Crainiceanu for provision of their R-scripts and helpful comments on the implementation of functional data analysis in accelerometry data. Furthermore, we would like to thank all subjects for their participation.

## Author contributions

Design of the original trials and securing of funding: AF, NK

Design of the analysis: TR, KN, UG

Data collection: SP, JL Data analysis: KN, TR

Statistical analysis: KN, TR

Paper writing: TR, KN, SP

All authors read and approved the final manuscript.

### Conflict of interest

None declared.

### Data sharing

Data and analysis scripts are available upon request at https://doi.org/10.5281/zenodo.3718578

## List of Abbreviations

BDI: Becks Depression Inventory
CI: Confidence interval
d-MEQ: Morningness-Eveningness-Questionnaire (german version)
ESS: Epworth Sleepiness Scale
FDA: Functional data analysis
FoSR: Function-on-scalar-regression
MCI: Mild cognitive impairment
MMSE: Mini-Mental-State-Examination
MRI: Magnetic Resonance Imaging
HOV: Healthy older volunteers
PANAS: Positive and Negative Affect Schedule
PSQI: Pittsburgh Sleep Quality Index
SMD: Standardized mean difference
STAIG-X1: State-Trait Anxiety Inventory (german version)
SVF: Stress-Verarbeitungs Fragebogen (translated: stress coping questionnaire)
tDCS: transcranial direct current stimulation
WHOQoL: World Health Organization Quality of Life

## Supporting information

**S1 Table. Inclusion and exclusion criteria**

**S2 Table. Neuropsychological test battery**

**S1 Appendix. Sleep and activity assessment**

**S2 Appendix. Statistical appendix**

**S1 Figure. Flow Chart**

**S2 Figure. Positioning of actigraph device**

## Notes

### Competing Interest Statement

The authors have declared no competing interest.

